# Prevalence and Clinical Correlates of Polycystic Ovarian Syndrome in Patients with Interstitial Cystitis/Bladder Pain Syndrome

**DOI:** 10.64898/2026.10.20.26366044

**Authors:** Hannah Ruetten, Kaylee Ferrara, Amr El Haraki, Robert Evans, Gopal Badlani, Stephen J. Walker

## Abstract

**Introduction and Hypothesis:** Polycystic Ovarian Syndrome (PCOS) occurs in 10% of women and hyperandrogenism is correlated with a symptom profile of pain and urinary dysfunction which overlaps with IC/BPS. In this study, we perform a cursory evaluation of the IC/BPS patient population to determine prevalence of PCOS and compare clinical features between patients with and without PCOS. We hypothesize that prevalence of PCOS in IC/BPS will be comparable to the general population and patients will have a unique symptom profile.

**Methods:** We performed a retrospective chart review of female patients (n=470) in our IC/BPS registry (IRB00018552) who underwent therapeutic hydrodistension and compared patients with PCOS diagnosis (n=15) to all other female patients (n=455) as well as an age, race, and parity matched control group (n=16). Univariate analysis was used to compare demographics and clinical features (Fisher exact test for categorical value comparison and Mann-Whitney U test for continuous variables). Variables significant on univariate analysis were then assessed with multiple logistic regression.

**Results:** We found that 3.19% of IC/BPS registry patients had a concurrent PCOS diagnosis. Patients were younger at initial IC/BPS diagnosis and had a higher BMI than non-PCOS patients. Patients with PCOS had similar IC/BPS features, symptom scores, and patient reported co-morbidities and non-urologic conditions to age, race, and parity match controls.

**Conclusions:** Further evaluation is needed of this IC/BPS subgroup to determine if low PCOS prevalence is due to a protective effect against IC/BPS and/or if hyperandrogenism contributes to the etiopathology of IC/BPS in patients with concurrent PCOS.

**Brief Summary:** Only 3.19% of IC/BPS patients receiving tertiary level urology care and hydrodistension treatment have concurrent PCOS. These patients are younger and have a higher BMI.

## Introduction

Interstitial cystitis/bladder pain syndrome (IC/BPS) is characterized by chronic pain arising from the bladder/pelvis associated with lower urinary tract symptoms (urgency, frequency and nocturia) [1–3]. Many patients experience localized suprapubic pain, but others have radiating pain to the urethra, vagina, and/or perineum. Patient symptoms are severe, significantly disrupt patients’ lives, and are associated with high rates of depression, anxiety, and low quality of life scores. This condition is life-long often with no current cure.

The etiology and pathophysiology of IC/BPS remains unclear despite decades of research, and the heterogeneous clinical presentation renders diagnosis and treatment a significant challenge. Comorbid pelvic conditions are a significant confounding factor in diagnosis and treatment as many co-occur at relatively high rates in IC/BPS patients [4]. Comorbid conditions such as pelvic floor dysfunction, vulvodynia, and endometriosis are often screened for during IC/BPS diagnosis and noted. However, little attention has been paid to concurrent polycystic ovarian syndrome (PCOS) despite its high prevalence in women.

PCOS occurs in 1 in 10 women of reproductive age [5]. Clinical/biochemical hyperandrogenism (elevated androgens) is diagnosed in 80% of PCOS patients and it is generally accepted that hyperandrogenism is the major driving factor behind hallmark signs and symptoms in PCOS, as well as other comorbidities found at high rates in these women [6]. Human clinical studies have reported a relationship between PCOS, hyperandrogenism, and lower urinary tract symptoms (LUTS) [7–9], including a significant positive correlation between serum testosterone levels and symptom and bother scores on the Pelvic Pain and Urinary Urgency/Frequency (PUF) Patient Symptom Scale. Based on PUF responses from 140 women with PCOS, patients’ hyperandrogenism may lead to more severe symptoms of dyspareunia, urgency, nocturia, and bladder/pelvic pain compared to the general population [9]. This symptom profile of bladder pain and urine storage dysfunction overlaps with interstitial cystitis/ bladder pain syndrome (IC/BPS) [1–3]. Therefore, in this study, we perform a cursory evaluation of the IC/BPS patient population with concurrent PCOS to determine: 1) prevalence of PCOS clinical diagnosis among IC/BPS patients, and 2) compare IC/BPS clinical features between patients with and without PCOS. Our overarching goal is to generate hypotheses for future prospective studies that can define distinct IC/BPS phenotypes that may share a similar targetable etiopathology.

## Materials and Methods

This retrospective cohort study involved a medical record review of previously collected data from consenting 18–80-year-old patients (IRB00018552) who were undergoing therapeutic hydrodistension (HOD) for IC/BPS. Demographic data, along with patient-reported history of co-morbidities and non-urologic syndromes (fibromyalgia, chronic pelvic pain, chronic fatigue syndrome, migraine, depression, panic disorder, general anxiety disorder, vulvodynia, pelvic floor dysfunction, endometriosis, sicca syndrome, POTS, IBS, allergies, and asthma), were recorded. Patients completed validated questionnaires, including the O’Leary-Sant Interstitial Cystitis Symptom and Problem Index (ICSI and ICPI), and the PUF patient symptom scale, before therapeutic HOD. Responses were excluded if patient provided written answers not listed on the survey or failed to respond to an item. An exception was made when patients recorded a value exceeding the maximum of the response scale; in these cases, the response was recorded to the highest permissible value. Patients were excluded from analysis if they were pregnant or had a history of radiation cystitis. At time of HOD (infusion into the bladder of sterile water for 5 minutes at a pressure of 100 cm H2O), additional clinical characteristics of IC/BPS such as presence of Hunner lesions and anesthetic bladder capacity were collected. Pathology reports for bladder mucosal biopsies taken at the time of HOD were also collected and analyzed for inflammation characteristics and mast cell count.

A manual medical record search for diagnosis of “PCOS”, “polycystic ovarian syndrome”, and/or “polycystic ovaries” was performed for all unique female IC/BPS patients meeting inclusion criteria.

An age, race, and parity matched cohort was established for a refined comparator group. Each PCOS patient was matched to a single age (+/-2 years), race, and parity matched control. When a race-matched patient of similar age and parity was not available in the cohort two patients were selected, one matching age and parity and one matching race.

Univariate analysis was used to compare demographics and clinical features between participants with and without co-occurring PCOS. The Fisher exact test was used for categorical value comparison, and the Mann-Whitney U test was used for continuous variables. P values were considered a trend at 0.05-0.1 and significant if ≤ 0.05. Significant differences on univariate analysis were then included in a subsequent multiple logistic regression. Statistical analyses were performed with Graph Pad Prism 10.2.3.

A manual search of all IC/BPS patient records from patients with concurrent PCOS was performed and any antiandrogen oral medications (Spironolactone, Finasteride, and/or Flutamide) were noted. Any information on androgen testing was also collected. No formal statistics were performed due to limited data available.

## Results

We screened 791 patient encounters and identified 470 patients meeting our inclusion criteria (Figure 1). Of the 470, 15 (3.19%) had a concurrent PCOS diagnosis.

**Figure 1.**
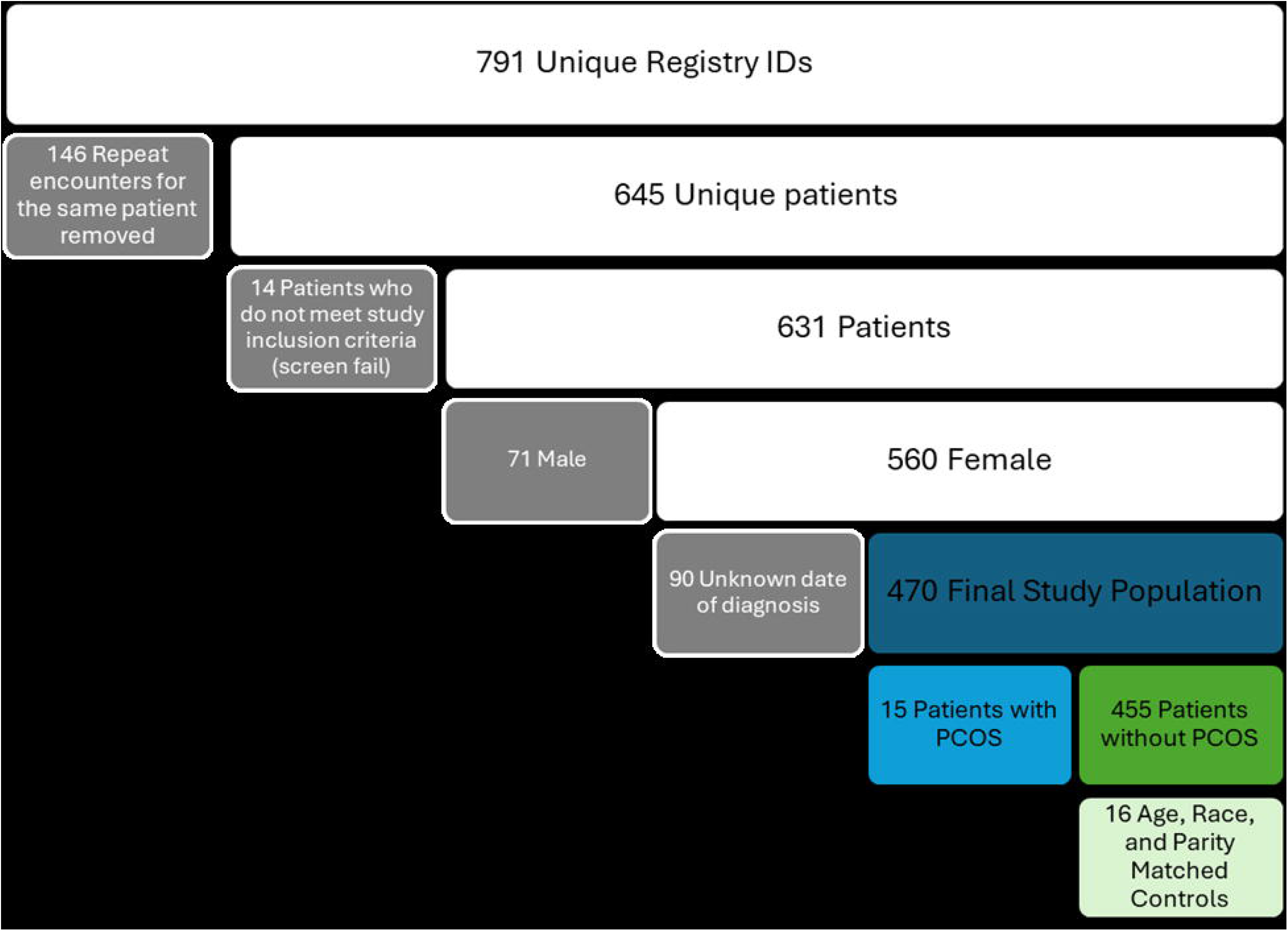
Patient selection from parent interstitial cystitis/bladder pain syndrome registry.

PCOS patient demographics were initially compared to the entire non-PCOS patient population. Patient average age was younger (25.80 vs 37.65 years, p=0.0028) at IC/BPS diagnosis. PCOS patients also had a higher average BMI (37.34 vs 29.17, p=0.0012) and there was a trend toward a difference in parity with 46.67% of the PCOS population being nulliparous versus 23.96% in the non-PCOS population (p=0.0741). Age at diagnosis and BMI were included in a multiple logistic regression analysis and younger age and higher BMI remained significantly associated with the PCOS population (Table 2).

Due to the low prevalence of PCOS, which limited the number of patients available for analysis, and marked differences in patient demographics, we established a non-PCOS control group matched for age, race, and parity for use in the remainder of study comparisons. There were no demographic differences between the PCOS population and the non-PCOS age, race, and parity matched control group. Detailed demographic information for each patient population can be found in Table 1.

**Table 1.** Polycystic ovarian syndrome (PCOS) and non-PCOS patient demographic information.

|  | All non-PCOS<br>n=455 | PCOS<br>n=15 | Age (A), Race<br>(R), and<br>Parity (P)<br>Matched<br>Control<br>n=16 | p-value<br>PCOS vs. All<br>non-PCOS | p-value<br>PCOS vs A,<br>R, and P<br>Matched<br>Control |
| --- | --- | --- | --- | --- | --- |
| Age at diagnosis | 37.65<br>(10-75) | 25.80<br>(13-43) | 26.88<br>(18-45) | 0.0028 | >0.9999 |
| Body mass index | 29.17<br>(14.91-54.90) | 37.34<br>(22.95-56.70) | 31.47<br>(17.30-46.00) | 0.0012 | 0.1879 |
| Parity |  |  |  | 0.0741 | 0.7982 |
| 0 | 109 (23.96%) | 7 (46.67%) | 9 (56.25%) |  |  |
| 1 | 82 (18.02%) | 4 (26.67%) | 4 (25.00%) |  |  |
| 2 | 155 (34.07%) | 1 (6.67%) | 2 (12.50%) |  |  |
| 3 | 62 (13.63%) | 1 (6.6+7%) | 1 (6.25%) |  |  |
| 4+ | 15 (3.30%) | 0 (0%) | 0 (0%) |  |  |
| Unknown | 32 (7.03%) | 2 (13.33%) | 0 (0%) |  |  |
| Race |  |  |  | 0.1106 | 0.3192 |
| Caucasian | 409 (89.89%) | 12 (80.00%) | 13 (81.25%) |  |  |
| African American | 35 (7.69%) | 2 (13.33%) | 2 (12.5%) |  |  |
| Oceanic/ Pacific | 2 (0.44%) | 1 (6.67%) | 1 (6.25%) |  |  |
| Islander | 4 (0.88%) | 0 (0%) | 0 (0%) |  |  |
| Hispanic | 5 (1.01%) | 0 (0%) | 0 (0%) |  |  |
| Other |  |  |  |  |  |

**Table 2.** Polycystic ovarian syndrome (PCOS) and non-PCOS patient demographic multiple logistic regression.

|  | Odds Ratio | Lower CI | Upper CI | p-value |
| --- | --- | --- | --- | --- |
| Age at Diagnosis | 1.075 | 1.008 | 1.166 | 0.0254 |
| Body mass index | 0.9129 | 0.8489 | 0.9790 | 0.0113 |

In addition, no IC/BPS characteristic differences, ICSI/ICPI score differences, PUF score differences, patient reported co-morbidities, or non-urologic syndromes differences were appreciated between the PCOS population and non-PCOS age, race, and parity matched controls (Supplemental Digital Content 1-4).

Pathology reports were read and assessed for mention of inflammation and mast cell count for all PCOS patients and non-PCOS age, race, and parity matched controls. A majority of reports diagnosed mucosal inflammation in both groups (66.67% in PCOS vs 84.62% in non-PCOS). Pathologists used a similar range of mast cells stains, and mast cell counts were similar between groups. Summarized pathology report information for each patient population can be found in Supplemental Digital Content 5.

Of the 15 patients with PCOS, only four had androgen testing performed and a report available in the electronic medical record system. Total testosterone values ranged from <10 to 128 ng/dL and only two of the four patients had a value above reference range. Unfortunately, all measurements were either years prior or after HOD (−1.5 to 8.2 years from HOD) so no correlations with IC/BPS features were performed.

Of the 15 patients diagnosed with PCOS, only one was receiving antiandrogen therapy (Spironolactone, Finasteride, and/or Flutamide) at the time of hydrodistension. They were receiving Spironolactone and had the highest total PUF score in the PCOS patient group (total score 31/35) as well as relatively high ICSI (14/19) and ICPI (14/16) scores. Another had notes of potential Spironolactone use around the time of hydrodistension, but a nurse practitioner noted that it was unclear whether the patient ever started taking the medication. A third patient began Spironolactone years after HOD.

## Discussion

We performed an initial exploratory study to determine prevalence and clinical features of IC/BPS patients with concurrent PCOS. Despite small numbers this study revealed a few findings warranting further attention and future study: 1) PCOS patient prevalence in IC/BPS is lower than in the general population, and 2) PCOS patients are younger and have an increased BMI compared to the general IC/BPS population.

PCOS is reported to occur in 10% of women of reproductive age [5], however in our IC/BPS population the prevalence was only 3.19%. This could be an artifact of our small sample size, a failure to capture history of non-urologic conditions at a tertiary level urology clinic, or a true underrepresentation of PCOS in the IC/BPS population. It is possible that concurrent PCOS prevalence is lower because PCOS may mask some features of IC/BPS that would lead a patient to seek urologic care. For example, patients with PCOS have increased pelvic floor muscle tone and maximum voluntary contraction [10] which could result in an increased ability to hold urine through an urge to void resulting in less urine leaking. They also may attribute pelvic pain to cystic ovaries rather than bladder pain, resulting in lower urology-related care seeking. Conversely, PCOS may confer a protective effect against IC/BPS due to a currently unknown mechanism. Further study is needed with increased patient numbers to determine if this reduced prevalence persists, as well as studies to investigate potential protective effects of PCOS against IC/BPS.

We also found that PCOS patients were younger at IC/BPS diagnosis and had an increased BMI compared to non-PCOS patients. The finding of a younger age at diagnosis was interesting as our group also found that patients with concurrent endometriosis were younger at IC/BPS diagnosis [11]. This finding may suggest that concurrent non-malignant gynecologic conditions predispose patients to the development of IC/BPS, potentially contributing to earlier symptom onset. The observed increase in BMI was not unexpected as elevated BMI is often associated with the metabolic syndrome that accompanies PCOS. However, given prior evidence demonstrating a high prevalence of metabolic syndrome in the broader IC/BPS population [12], it was somewhat surprising that PCOS patients exhibited an even higher BMI. Future studies are needed to determine whether a distinct subgroup of IC/BPS patients characterized by “early onset” of symptoms exists as well as to clarify the contribution of metabolic syndrome to the development and severity IC/BPS symptoms.

In a prior study PCOS patients’ serum testosterone level had a significant positive correlation with dyspareunia, urgency, nocturia, and bladder/pelvic pain [9]. In their cohort, PUF total scores ranged from 0-23 with an average score of 4.6 [9]. In contrast, PCOS patients in our study demonstrated markedly higher PUF total scores ranging from 20-31 with an average score of 25. These findings confirm that while IC/BPS symptoms occur in a subset of patients with PCOS, not all individuals with PCOS experience IC/BPS. However, this previous study’s finding that serum testosterone has a significant positive correlation with more severe symptom scores on the PUF survey [9] could mean that a subset of PCOS patients, who experience high serum testosterone levels, could develop IC/BPS-like symptoms related to their high testosterone. Unfortunately, due to the retrospective nature of this study we were unable to capture serum androgen levels at time of HOD. A prospective cohort study that includes serum hormone assessment would provide necessary correlation data to further implicate androgens in the pathogenesis of IC/BPS and urinary dysfunction.

There are several limitations of this study. The first is that PCOS patients available for assessment were determined by medical chart notation of “PCOS”, “polycystic ovarian syndrome”, and/or “polycystic ovaries” rather than Rotterdam criteria. It is possible that some of the patients included in our cohort do not meet Rotterdam criteria. It is also possible that some IC/BPS patients have an undiagnosed co-occurring PCOS. In addition, it is possible that some non-PCOS controls do in fact have PCOS. In the future, a study that includes measurement of serum hormones, ultrasonographic assessment of ovaries, and assessment of menstrual cycle would provide a more rigorous study design fitting with current PCOS diagnostic guidelines. However, many studies use patient reported diagnosis of co-morbid conditions for assessment and our group definitions are of equivalent rigor to a patient reported grouping. Second, due to the low prevalence of PCOS in our registry cohort, we had low numbers of patients for assessment. We used an age, race, and parity matched control group to control for confounding variables but it is possible that due to low numbers we have a higher degree of type I and type II error in our statistical interpretation.

PCOS patients comprise a younger IC/BPS patient population with higher BMI. This study provides initial data to build on in future studies focused on age, BMI, PCOS, and serum testosterone levels in IC/BPS. IC/BPS is a complex disease to diagnose and treat. Only by parsing out the unique patient subpopulations that comprise this heterogenous patient population can we hope to better understand IC/BPS etiopathology.

## Supporting information

Supplemental Digital Content 5

Supplemental Digital Content 1

Supplemental Digital Content 2

Supplemental Digital Content 3

Supplemental Digital Content 4

## Data Availability

All data produced in the present work are contained in the manuscript.

## Financial Disclaimers/Conflict of Interest Statement

We declare that we have no conflict of interest. The funders had no role in the design of the study; in the collection, analyses, or interpretation of data; in the writing of the manuscript; or in the decision to publish the results.

## Funding

This study was partially funded by R21DK106554-01 (SJW) and 1R01DK124599-01 (SJW). Ruetten was supported by T32 OD010957, L70 DK140978, and K99 DK149117-01.

## Research Ethics Approval

This study involves human participants, was conducted in accordance with the Declaration of Helsinki, and approved by the Institutional Review Board of Wake Forest Baptist Hospital (protocol code IRB00018552).

## Informed Consent Statement

Informed consent was obtained from all subjects involved in the study.

## Acknowledgments

The authors thank the study participants for making this study possible.

## Author Contributions (Protocol/project development, Data collection or management, Data analysis, Manuscript writing/editing, Other (please specify briefly using 1 to 5 words))

H Ruetten: Project development, data collection, data analysis, manuscript writing

K Ferrara: Data collection

A El Haraki: Manuscript editing, Other (IC/BPS patient registry development) R Evans: Other (IC/BPS patient registry development)

G Badlani: Other (IC/BPS patient registry development)

SJ Walker: Manuscript editing, Other (IC/BPS patient registry development/management)

## Supplemental Digital Content

Supplemental Digital Content 1.pdf

Supplemental Digital Content 2.pdf

Supplemental Digital Content 3.pdf

Supplemental Digital Content 4.pdf

Supplemental Digital Content 5.pdf

## Notes

### Competing Interest Statement

The authors have declared no competing interest.

### Author Declarations

IRB of Wake Forest University gave ethical approval for this work.

