## Supplemental Digital Content 5 for "Prevalence and Clinical Correlates of Polycystic Ovarian Syndrome in Patients with Interstitial Cystitis/Bladder Pain Syndrome"

**Polycystic ovarian syndrome (PCOS) and non-PCOS patient pathology report findings.**

|  | PCOS  n=9/15 | Age (A), Race (R), and Parity (P) Matched Control  n=13/16 | p-value |
| --- | --- | --- | --- |
| Inflammation | 6/9 (66.67%) | 11/13 (84.62%) | 0.6090 |
| Mast Cell Stain  C-kit  CD117  MCT | 2/8 (25.00%)  1/8 (12.50%)  5/8 (62.50%) | 3/11 (27.27%)  0/11 (0%)  8/11 (72.73%) | 0.7730 |
| Mast Cell Count | 15.31 (2-30)  n=8 | 24.18 (5-45)  n=11 | 0.1635 |
