## Supplemental Digital Content 1 for "Prevalence and Clinical Correlates of Polycystic Ovarian Syndrome in Patients with Interstitial Cystitis/Bladder Pain Syndrome"

**Polycystic ovarian syndrome (PCOS) and non-PCOS patient initial cystitis/bladder pain syndrome (IC/BPS) characteristics.**

|  | PCOS  n=15 | Age (A), Race (R), and Parity (P) Matched Control  n=16 | p-value |
| --- | --- | --- | --- |
| Age at diagnosis of IC/BPS in years (range) | 25.80  (13-43) | 26.88  (18-45) | >0.9999 |
| Bladder capacity in mL (range) | 853.3  (450-1400) | 859.4  (400-1300) | >0.9999 |
| Low bladder capacity (%) | 1 (6.67%) | 2 (12.50%) | >0.9999 |
| Hunner lesion positive (%) | 0 (0%)  n=14/15 | 2 (12.50%) | 0.4851 |
| Undergone Cystectomy (%) | 0 (0%)  n=11/15 | 0 (0%)  n=15/16 | >0.9999 |
