## Supplemental Digital Content 2 for "Prevalence and Clinical Correlates of Polycystic Ovarian Syndrome in Patients with Interstitial Cystitis/Bladder Pain Syndrome"

**Polycystic ovarian syndrome (PCOS) and non-PCOS patient ICSI and ICPI responses.**

|  | PCOS  n=15 | Age (A), Race (R), and Parity (P) Matched Control  n=16 | p-value |
| --- | --- | --- | --- |
| ICSI score (range)  Urgency  0  1  2  3  4  5  Frequency  0  1  2  3  4  5  Nocturia  0  1  2  3  4  5  Pain/Burning  0  1  2  3  4 | 12.91 (9-17)  n=11/15  0 (0%)  0 (0%)  2 (18.18%)  4 (36.36%)  4 (36.36%)  1 (9.09%)  1 (9.09%)  0 (0%)  0 (0%)  1 (9.09%)  6 (54.55%)  3 (27.27%)  1 (9.09%)  0 (0%)  4 (36.36%)  2 (18.18%)  1 (9.09%)  3 (27.27%)  0 (0%)  1 (9.09%)  4 (36.36%)  4 (36.36%)  2 (18.18%) | 14.13 (10-17)  n=15/16  0 (0%)  2 (13.33%)  0 (0%)  3 (20.00%)  9 (60.00%)  1 (6.67%)  0 (0%)  1 (6.67%)  0 (0%)  1 (6.67%)  7 (46.67%)  6 (40.00%)  0 (0%)  1 (6.67%)  4 (26.67%)  7 (46.67%)  1 (6.67%)  2 (13.33%)  0 (0%)  0 (0%)  2 (13.33%)  4 (26.67%)  9 (60.00%) | 0.7649  0.2620  0.8181  0.5274  0.1347 |
| ICPI score (range)  Urgency  0  1  2  3  4  Frequency  0  1  2  3  4  Nocturia  0  1  2  3  4  Pain/Burning  0  1  2  3  4 | 13.36 (11-15)  n=11/15  0 (0%)  0 (0%)  1 (9.09%)  6 (54.55%)  4 (36.36%)  0 (0%)  0 (0%)  0 (0%)  6 (54.55%)  5 (45.46%)  1 (9.09%)  0 (0%)  1 (9.09%)  4 (36.36%)  5 (45.46%)  0 (0%)  0 (0%)  2 (18.18%)  0 (0%)  9 (81.82%) | 13.13 (9-16)  n=15/16  0 (0%)  0 (0%)  5 (33.33%)  6 (40.00%)  4 (26.67%)  0 (0%)  0 (0%)  1 (6.67%)  8 (53.33%)  6 (40.00%)  0 (0%)  2 (13.33%)  1 (6.67%)  6 (40.00%)  6 (40.00%)  0 (0%)  0 (0%)  0 (0%)  3 (20.00%)  12 (80.00%) | >0.9999  0.4128  >0.9999  0.7532  0.1015 |
