## Supplemental Digital Content 3 for "Prevalence and Clinical Correlates of Polycystic Ovarian Syndrome in Patients with Interstitial Cystitis/Bladder Pain Syndrome"

**Polycystic ovarian syndrome (PCOS) and non-PCOS patient PUF responses.**

|  | PCOS  n=15 | Age (A), Race (R), and Parity (P) Matched Control  n=16 | p-value |
| --- | --- | --- | --- |
| PUF score | 25.00 (20-31)  n=10/15 | 23.86 (17-33)  n=14/16 | >0.9999 |
| PUF score- Symptom  Frequency  0  1  2  3  4  Nocturia  0  1  2  3  4  Pain w/ Sex  0  1  2  3  Bladder/Pelvis  Pain Frequency  0  1  2  3  Pain Severity  1  2  3  Urgency  0  1  2  3  Urgency Severity  1  2  3 | 16.20 (14-19)  n=10/15  0 (0%)  2 (20.00%)  4 (40.00%)  2 (20.00%)  2 (20.00%)  1 (10.00%)  1 (10.00%)  3 (30.00%)  1 (10.00%)  4 (40.00%)  1/9 (11.11%)  4/9 (44.44%)  0/9 (0%)  4/9 (44.44%)  0 (0%)  1 (10.00%)  2 (20.00%)  7 (70.00%)  2 (20.00%)  5 (50.00%)  3 (30.00%)  0 (0%)  0 (0%)  6 (60.00%)  4 (40.00%)  0 (0%)  5 (50.00%)  5 (50.00%) | 15.29 (10-21)  n=14/16  1 (7.14%)  6 (42.86%)  4 (28.57%)  2 (14.29%)  1 (7.14%)  0 0%)  1 (7.14%)  5 (35.71%)  4 (28.57%)  4 (28.57%)  1 (7.14%)  3 (21.43%)  6 (42.86%)  4 (28.57%)  0 (0%)  1 (7.14%)  4 (28.57%)  9 (64.29%)  1 (7.14%)  6 (42.86%)  7 (50.00%)  1 (7.14%)  4 (28.57%)  5 (35.71%)  4 (28.57%)  3 (21.43%)  8 (57.14%)  3 (21.43%) | >0.9999  0.6678  0.8061  0.1052  0.8401  0.6184  0.2052  0.2099 |
| PUF score- Bother  Nocturia  0  1  2  3  Pain w/ Sex  0  1  2  3  Pain  0  1  2  3  Urgency  0  1  2  3 | 8.80 (5-12)  n=10/15  1 (10.00%)  1 (10.00%)  5 (50.00%)  3 (30.00%)  1/9 (11.11%)  2/9 (22.22%)  4/9 (44.44%)  2/9 (22.22%)  0 (0%)  2 (20.00%)  1 (10.00%)  7 (70.00%)  0 (0%)  0 (0%)  3 (30.00%)  7 (70.00%) | 8.57 (6-12)  n=14/16  0 (0%)  4 (28.57%)  2 (14.29%)  8 (57.14%)  2 (14.29%)  6 (42.86%)  4 (28.57%)  2 (14.29%)  0 (0%)  1 (7.14%)  2 (14.29%)  11 (78.57%)  1 (7.14%)  3 (21.43%)  3 (21.43%)  7 (50.00%) | >0.9999  0.1345  0.7697  0.7992  0.4772 |
