## Supplemental Digital Content 4 for "Prevalence and Clinical Correlates of Polycystic Ovarian Syndrome in Patients with Interstitial Cystitis/Bladder Pain Syndrome"

**Polycystic ovarian syndrome (PCOS) and non-PCOS patient-reported history of co-morbidities and non-urologic syndromes.**

|  | PCOS  n=15 | Age (A), Race (R), and Parity (P) Matched Control  n=16 | p-value  PCOS vs A, R, and P Matched Control |
| --- | --- | --- | --- |
| Fibromyalgia | 5/9 (55.56%) | 4/10 (40.00%) | 0.6563 |
| Chronic Pelvic Pain | 4/7 (57.14%) | 3/10 (30.00%) | 0.3500 |
| Chronic Fatigue | 1/8 (12.50%) | 3/7 (30.00%) | 0.5882 |
| Migraine | 5/8 (62.50%) | 4/10 (40.00%) | 0.6372 |
| Depression | 4/7 (57.14%) | 5/10 (50.00%) | >0.9999 |
| Panic Disorder | 4/7 (57.14%) | 4/11 (36.36%) | 0.6305 |
| General Anxiety Disorder | 5/6 (83.33%) | 6/8 (75.00%) | >0.9999 |
| Vulvodynia | 1/7 (14.29%) | 3/10 (30.00%) | 0.6029 |
| Pelvic Floor Dysfunction | 6/9 (66.67%) | 4/10 (40.00%) | 0.3698 |
| Endometriosis | 3/9 (33.33%) | 4/10 (40.00%) | >0.9999 |
| Sicca Syndrome | 0/7 (0%) | 0/10 (0%) | >0.9999 |
| POTS | 0/6 (0%) | 1/8 (12.5%) | >0.9999 |
| IBS | 4/8 (50.00%) | 5/10 (50.00%) | >0.9999 |
| Allergies  Seasonal  Medication  Food | 7/9 (77.78%)  1/4 (25.00%)  1/4 (25.00%)  0/4 (25.00%) | 7/11 (63.64%)  2/7 (28.57%)  3/7 (42.86%)  0/7 (0%) | 0.6424  >0.9999  >0.9999  >0.9999 |
| Asthma | 4/8 (50.00%) | 4/10 (40.00%) | >0.9999 |
